# Outbreak-associated *Salmonella* Baildon found in wastewater demonstrates how sewage monitoring can supplement traditional disease surveillance

**DOI:** 10.1101/2024.07.03.24308516

**Authors:** Nkuchia M. M’ikanatha, Zoe S. Goldblum, Nicholas Cesari, Erin M. Nawrocki, Yezhi Fu, Jasna Kovac, Edward G. Dudley

**Affiliations:** Division of Infectious Disease Epidemiology, Pennsylvania Department of Health, Harrisburg, PA, USA; Center for Clinical Epidemiology and Biostatistics, Perelman School of Medicine, University of Pennsylvania, Philadelphia, PA, USA; Department of Food Science, The Pennsylvania State University, University Park, PA, USA; The E. coli Reference Center, The Pennsylvania State University, University Park, PA, USA

**Author notes:** Corresponding Author and Reprints: Nkuchia M. M’ikanatha, DrPH, MPH; Division of Infectious Disease Epidemiology, Pennsylvania Department of Health, Harrisburg, PA 17120. Department of Microbiology and Molecular Genetics, University of Pittsburgh, Pittsburgh, Pennsylvania (Erin M. Nawrocki). School of Agriculture, Shenzhen Campus of Sun Yat-sen University, Shenzhen, China (Yezhi Fu).

**Keywords:** *Salmonella*, communicable disease, food safety, foodborne infections, disease outbreaks, foodborne pathogens, domestic wastewater surveillance, epidemiological monitoring

## Abstract

Non-typhoidal *Salmonella* is a common cause of gastroenteritis worldwide, but current non-typhoidal *Salmonella* surveillance is suboptimal. Here we evaluated the utility of wastewater monitoring to enhance surveillance for this foodborne pathogen. In June 2022, we tested composite raw sewage collected twice a week from two treatment plants in central Pennsylvania for non-typhoidal *Salmonella* and characterized isolates using whole genome sequencing. We recovered 43 *Salmonella* isolates from wastewater samples, differentiated by genomic analysis into seven serovars: 16 Panama (37.2%), 9 Senftenberg (20.9%), and 8 Baildon (18.6%), 3 or fewer for four other serovars. We assessed genetic relatedness and epidemiologic links between non-typhoidal *Salmonella* serovars from wastewater and isolates from patients with salmonellosis. All *S.* Baildon serovars from wastewater were genetically and epidemiologically associated with a known salmonellosis outbreak. *S.* Baildon from wastewater and 42 outbreak-related isolates in the national outbreak detection database had the same core genome multilocus sequence typing and code differed by no or one single polynucleotide polymorphisms. One of the 42 outbreak-related isolates was obtained from a patient residing in the wastewater sample collection catchment area, which serves approximately 17000 people. *S.* Baildon is a rare serovar (reported in <1% cases nationally over five years). Our study underscores the value of monitoring sewage from a defined population to supplement traditional surveillance methods for evidence of *Salmonella* infections and to determine the extent of outbreaks.

**Importance statement:** During the COVID-19 pandemic, surveillance for SARS-CoV-2 in wastewater was a powerful tool for assessing community burden days ahead of traditional reporting. Here, we show that domestic sewage monitoring is useful for surveillance for a foodborne pathogen. *Salmonella enterica* was detected in samples from two wastewater treatment plants in central Pennsylvania during June 2022. Using whole genome sequencing, we demonstrated that isolates of variant *S*. Baildon clustered with those from an outbreak that occurred in a similar time frame. Case reports were primarily from Pennsylvania, and one individual lived within the treatment center catchment area. This study provides support for the utility of domestic sewage surveillance in assisting public health agencies identify communities impacted by infectious diseases.

## Introduction

Non-typhoidal *Salmonella* is a common cause of gastroenteritis worldwide. Current non-typhoidal *Salmonella* surveillance systems are based on reporting by healthcare professionals and clinical laboratories (1–3). Although there are variations across jurisdictions, the sensitivity and timeliness of these systems to detect outbreaks is limited by under-reporting, incomplete case reports, and resource constraints (4). Although an estimated 1.03 million cases of salmonellosis occur annually in the United States associated with 74,000 physician visits, 19,000 hospitalizations, 378 deaths, and $4.14 billion in direct and indirect costs (https://www.ers.usda.gov/data-products/cost-estimates-of-foodborne-illnesses/), in 2023, only 52,575 salmonellosis cases were reported by all jurisdictions in the United States through the National Notifiable Diseases Surveillance System (https://wonder.cdc.gov/nndss/static/2023/52/2023-52-table1122.html),

For diseases such as salmonellosis and COVID-19, unrecognized cases can amplify transmission (5–6). Evidence of infection can be detected in stool samples from approximately 90% of individuals infected with *Salmonella* and 40% of those with SARS-CoV2 (7–8). Identification and quantification of SARS-CoV-2 RNA in untreated influents from domestic wastewater treatment plants augmented traditional disease reporting during the pandemic and is now a major component of the COVID-19 surveillance in the United States (9). Wastewater-based surveillance detects SARS-CoV-2 RNA before the peak of clinical cases and has been shown to identify variants of concern prior to presentation in patients (10–11). Emergency appropriations in 2021 expanded wastewater-based surveillance efforts for COVID-19 through the US Food and Drug Administration GenomeTrakr program which has been focused on foodborne pathogens (12).

We participated in the wastewater monitoring for SARS-CoV-2 variants as part of the GenomeTrakr network laboratories and reasoned that the same monitoring could be useful for non-typhoidal *Salmonella* surveillance. The Pennsylvania Department of Health Bureau of Laboratories performs whole genome sequencing of non-typhoidal *Salmonella* found in clinical samples and uploads data to the Centers for Disease Control and Prevention (CDC) PulseNet program to facilitate outbreak investigations (13). Diagnostic laboratories send presumptive *Salmonella* isolates in compliance with communicable disease requirements, which also mandates the reporting of detailed patient information, including demographic characteristics, via the secure Pennsylvania National Electronic Disease Reporting system (PA-NEDSS) (14). Investigations of individual salmonellosis cases and outbreaks are documented in PA-NEDSS (15). Additionally, Pennsylvania sends some isolates from patients and all non-typhoidal *Salmonella* isolates found in retail food sources to the National Antimicrobial Resistance Monitoring System (NARMS) laboratories at the CDC and the Food and Drug Administration (FDA), respectively (16).

By leveraging the SARS-CoV-2 variants project and the existing state and national public health systems for salmonellosis (13–16), we conducted a study to assess whether wastewater-based surveillance could enhance surveillance for non-typhoidal *Salmonella*. We hypothesized that *Salmonella* strains linked to recent outbreaks would be detected in domestic sewage.

## Materials and Methods

### Catchment area population and characteristics

Samples were collected from two wastewater treatment facilities in central Pennsylvania. A wastewater treatment plant manager in each facility completed a brief survey that included information on the number of customers and presence of institutional residential facilities such as prisons within the catchment area. We used the household equivalent (customers served) combined with United States Census Bureau data to estimate the total population of each catchment area (https://www.census.gov/quickfacts/fact/table/PA/HSG010222). Twice weekly during June 13-29, 2022, a plant operator at each facility collected a 24-hour composite untreated sewage sample upstream of the primary clarifier following the United States Environmental Protection Agency protocol for sample collection (https://www.epa.gov/quality/procedures-collecting-wastewater-samples). Samples were stored at 4 °C in the dark for less than 48 h prior to being transported on ice to the laboratory for processing.

### Detection of Salmonella in raw wastewater

We followed standardized procedures for isolation of *Salmonella* (17–18). Briefly, we transferred 120 ml into a bottle, centrifuged at 5,000 x *g* for 20 min at 4 °C, filtered the supernatant through a 0.45-µm membrane and added the filter to 40 mL of buffered peptone water supplemented with 20 mg/L novobiocin. Broths were incubated at 35 °C for 24 h. Presumptive *Salmonella* colonies were sub-cultured in tetrathionate and Rappaport–Vassiliadis broths at 42 °C for 24 h. Aliquots of 10 µL of each enriched culture were then plated on Xylose Lysine Deoxycholate (XLD) and Hektoen Enteric (HE) agar. We selected eight presumptive *Salmonell*a colonies from each XLD and HE plate and were screened for *invA* by PCR (18). Forty-three were confirmed to be *Salmonella*, and thus sequenced (Supplemental Table 1).

### Whole genome sequencing and cluster detection

We extracted DNA from *Salmonella* isolates with the Qiagen DNeasy blood and tissue kit, and libraries were prepared using the Nextera XT DNA library kit. Libraries were sequenced using an Illumina MiSeq using the manufacturers reagent kits (V3) with 500 (2 x 250) cycles. We uploaded the short-read sequences to the National Center for Biotechnology Information (NCBI), and the NCBI Pathogen Detection Isolate Browser (NCBI-PD) automated bionformatics pipeline was used to process sequences and create phylogenetic single nucleotide polymorphism (SNP) trees (https://www.ncbi.nlm.nih.gov/pathogens/pathogens_help/#isolates-browser). The SeqSero2 application within the NCBI-PD pipeline was used to identify molecular serotypes (19). Additionally, we used the NCBI-PD pipeline-AMRFinderPlus to identify antimicrobial resistance gene sequences and mutations that matched with those in the Reference Gene Catalog (20).

### Verification of clusters and epidemiological investigations

We used the CDC Systems Enteric Disease Response, Investigation, and Coordination (SEDRIC) platform to verify clustering of isolates from clinical and wastewater sources (21). SEDRIC analyses are based on core genome multilocus sequence typing, a gene-by-gene comparison approach used for outbreak cluster detection. We considered isolates from clinical and wastewater sources related if they differed by ≤10 alleles (22). SEDRIC enabled us to pinpoint each isolate by geographic location, temporality, allele code, and associated multi-state outbreak code, if any. For related isolates from Pennsylvania, we reviewed original records on the state-based surveillance system. We also examined summaries of outbreaks including demographic and epidemiological characteristics of the case patients. To assess the potential for under-reporting, we examined salmonellosis cases reported from the catchment area during 2019-2022. We reviewed state and national surveillance systems to identify serovars associated with salmonellosis.

### Evolutionary analysis by the maximum likelihood method

We analyzed the genetic relatedness of isolates from clinical and wastewater sources using the FDA Center for Food Safety and Applied Nutrition (CFSAN) SNP Pipeline on the GalaxyTrakr website as previously described (23). The generated output snp_profile.fasta file was used to build a maximum likelihood tree using MEGA11 (Molecular Evolutionary Genetic Analysis) application (25).

### Ethical considerations

Access to data on clinical *Salmonella* isolates was approved by CDC NARMS state-based program activities.

## Results

### Catchment area characteristics

There were 5,600 and 1,500 equivalent households served by wastewater treatment plants A and B, respectively. The average persons per household in the state in 2022 was 2.42 (https://www.census.gov/quickfacts/fact/table/PA/HSG010222). Thus, the total population in the catchment area of the two Pennsylvania wastewater treatment plants from which samples were collected was estimated at 17,182. Facilities reported in the survey that they received domestic sewage only, without any storm water runoff or environmental runoff such as that from farm operations or poultry processing plants.

### Salmonella serovars isolated from wastewater

*Salmonella* was found in 9 of the 12 wastewater samples (75%) that were collected from the Pennsylvania plants in June 2022; 43 *Salmonella* isolates were recovered. As previously reported (26), based on whole genome assemblies, these isolates were from seven serovars: 16 were Panama (37.2%), 9 were Senftenberg (20.9%), and 8 were Baildon (18.6%). Three other serovars had 3 or fewer isolates (Fig. 2). There were no clinically relevant antimicrobial resistance genes or resistance-conferring SNPs identified in any of the isolates.

All eight (100%) *S.* Baildon isolates from domestic sewage samples were separated by 0 or 1 SNPs from 46 clinical isolates identified in SEDRIC (Fig. 3), and all were assigned the same allele code in the national outbreak database. Forty-four (95.7%) of the clinical isolates originated from Pennsylvania, and two (SRR19419658 and SRR19512122) were from nearby states (West Virginia and Virginia) (Supplemental Table 2). One case of salmonellosis associated with *S.* Baildon was reported in a zip code within the catchment area (Fig. 1).

**Figure 1.**
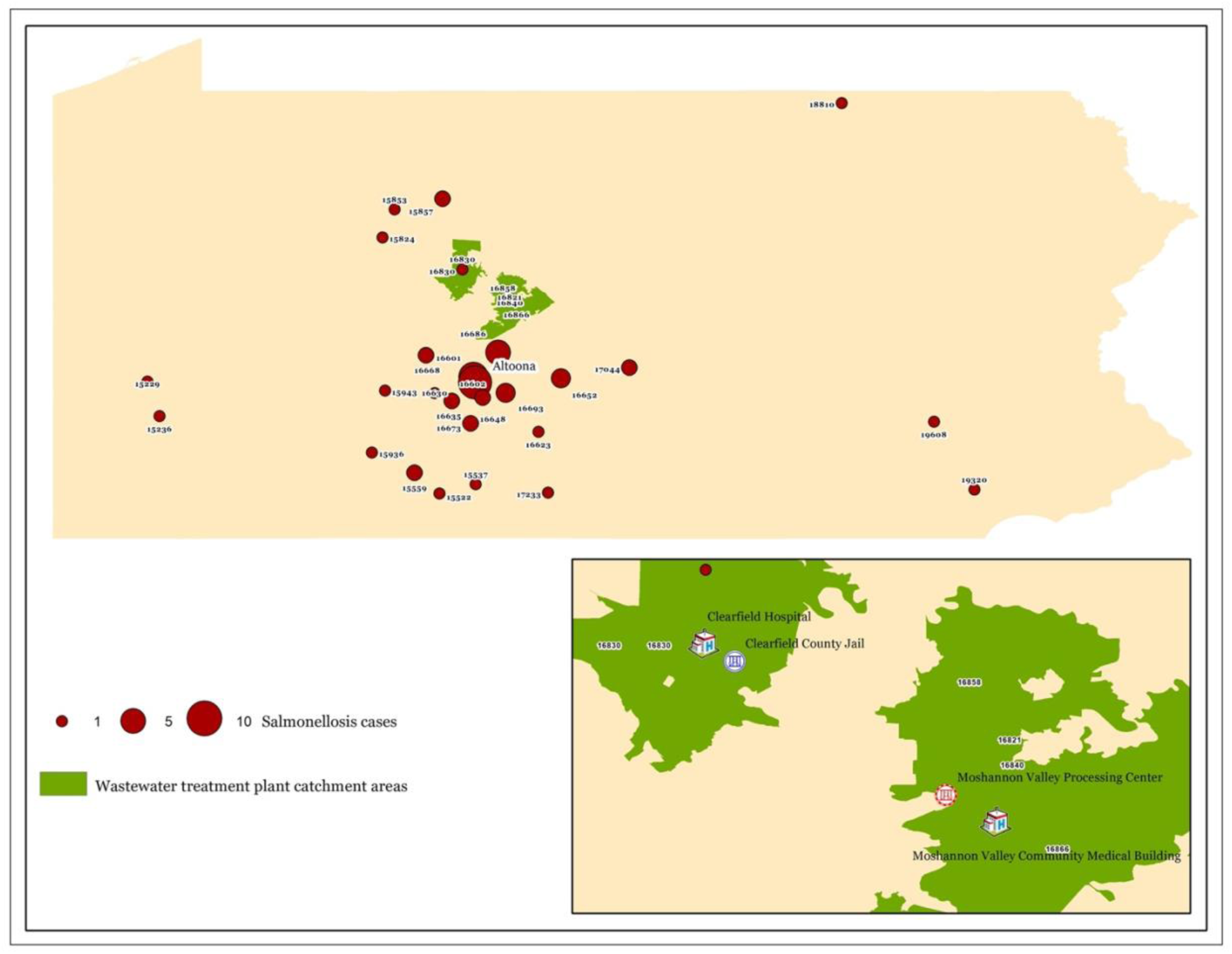
Locations of domestic sewage treatment plant catchment areas (green) and locations of zip codes where clinical cases were reported with numbers of cases indicated by sizes of the red dots. Also indicated are the location of the Clearfield County Jail, and Moshannon Valley Processing Center, which in addition to community households are included in the catchment area.

**Figure 2.**
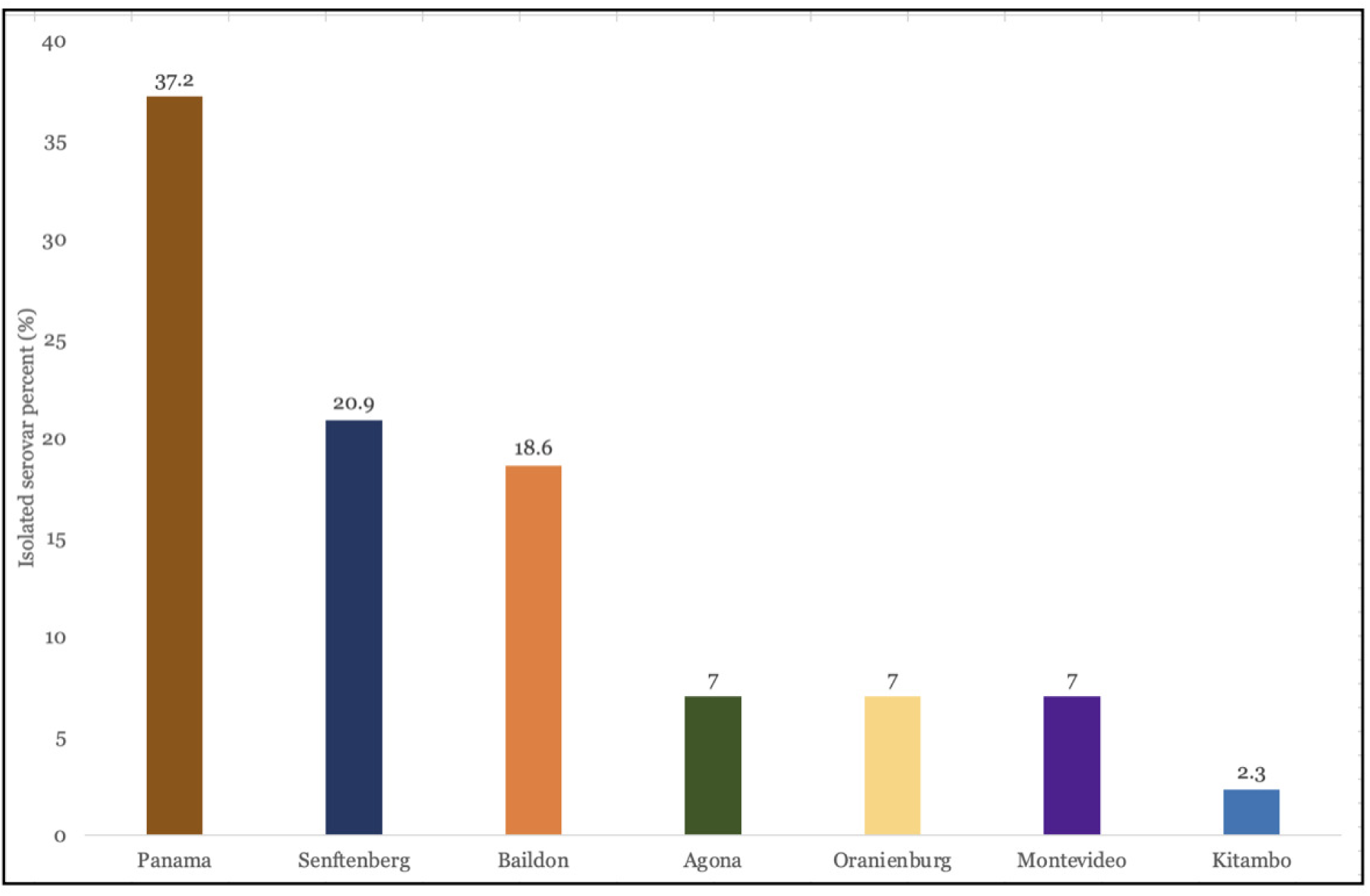
Distribution of non-typhoidal *Salmonella* serotypes identified among the 43 isolates collected during June 2022. The catchment area served by two wastewater treatment plants in central Pennsylvania served a total population of approximately 17,182.

**Figure 3.**
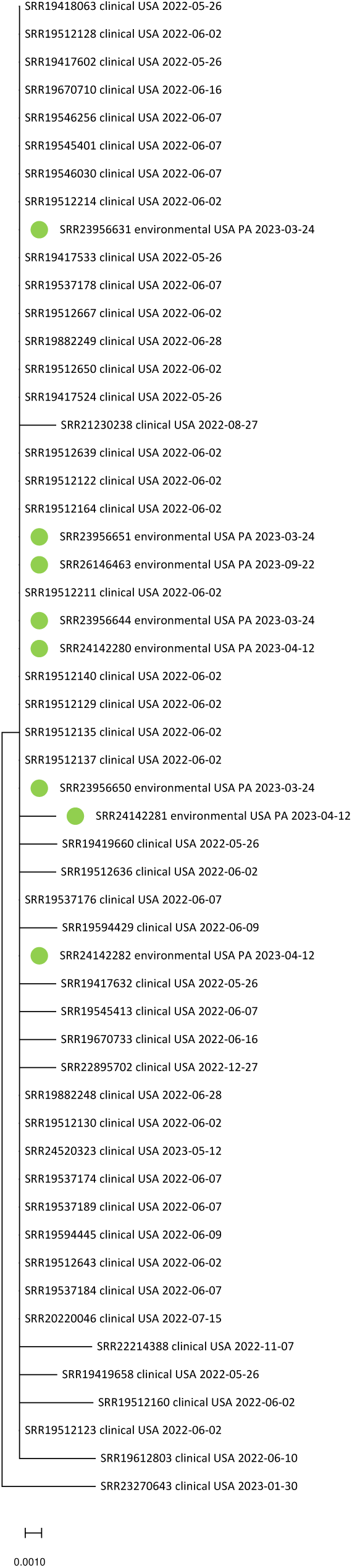
Genetic relatedness of *S*. Baildon isolates (n=8) from sewage sources (marked green) and clinical sources (n=46). Isolates from sewage samples were separated by 0 or 1 single nucleotide polymorphisms from those linked to salmonellosis. The tree is drawn to scale, with branch lengths measured in the number of substitutions per site. Evolutionary analyses were conducted in MEGA11 (30).

### Epidemiological investigation of genetically related S. Baildon isolates from wastewater and clinical sources

Forty-two (91.3%) of the 46 clinical isolates were traced to a salmonellosis outbreak that occurred May 3-12, 2022 in Pennsylvania, based on the case confirmation. The outbreak was epidemiologically linked to consumption of contaminated tomato and iceberg salad (odds ratio, 7.25, 95% confidence interval (1.98-26.54); *p*=0.003). The food was consumed starting on May 3, 2022 at a chain restaurant in Altoona, Pennsylvania, within an hour’s driving distance from the wastewater catchment areas (Fig. 1).

### S. Baildon reported in the catchment areas, state and national surveillance systems

Lastly, we investigated the reported prevalence of salmonellosis and *S*. Baildon on a local, state and national level. Our review of state-based surveillance found 11 salmonellosis cases were reported from the catchment during 2019-2023. Further examination of state and national surveillance systems showed that during 2019-2023, *S.* Baildon infections were associated with 53 of the 8,143 cases of salmonellosis reported in Pennsylvania (0.7%) and in 650 of 330,172 cases reported in the United States (0.2%) (Table 1).

**Table 1.**
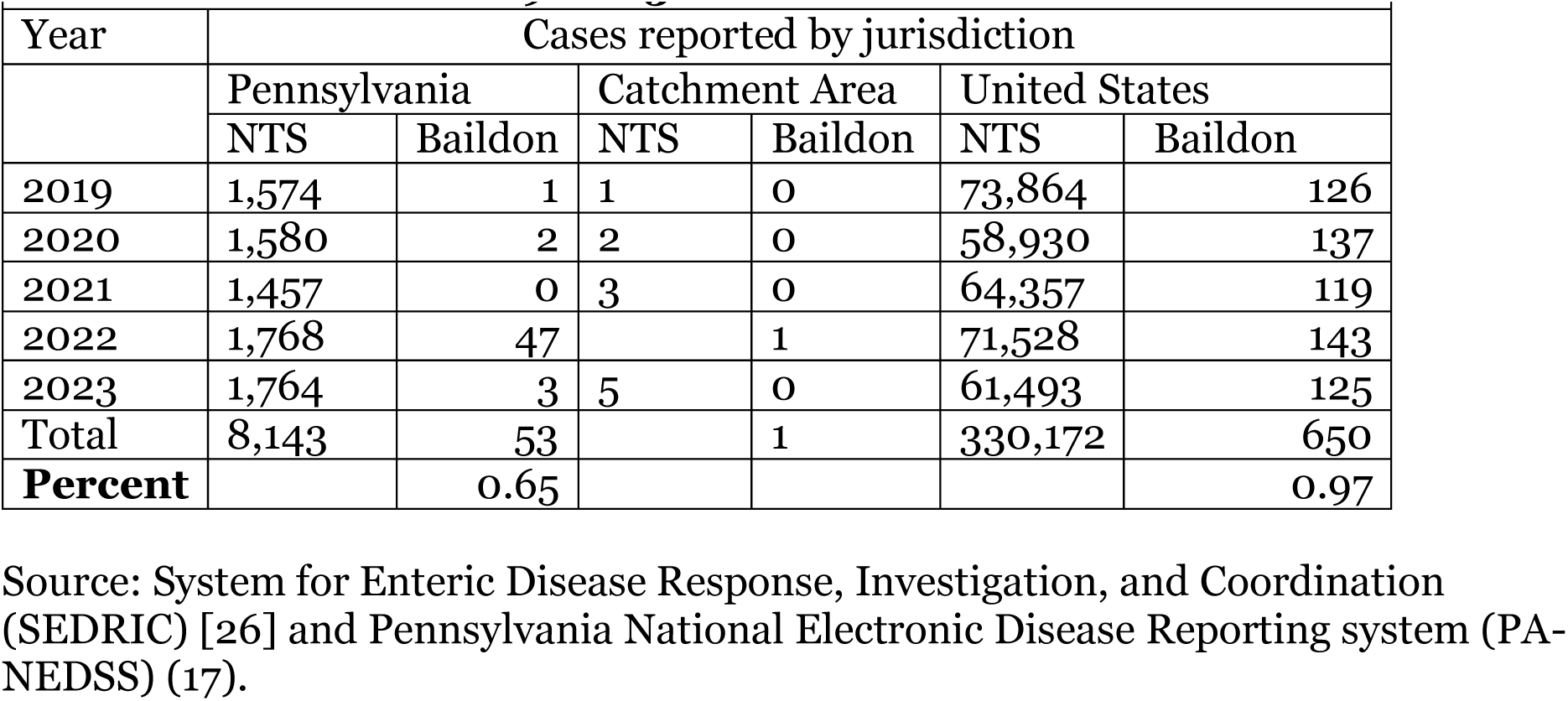
Prevalence of *Salmonella* Baildon in Pennsylvania and United States—2019-2023.

## Discussion

In June 2022, we tested untreated domestic wastewater from two facilities in central Pennsylvania that serve about 17,000 residents for non-typhoidal *Salmonella*. We isolated and sequenced 42 non-typhoidal *Salmonella* strains that were differentiated into seven serotypes. Isolates from two serovars, *S.* Baildon, as discussed here, and previously discussed *S.* Senftenberg (26), were genetically matched to clinical cases from the same time period. Infections associated with the two outbreaks were reported to public health authorities either concurrently with or immediate after we had detected *Salmonella* in wastewater samples from the catchment area. This *Salmonella* serotype is rare: *S*. Baildon accounted for less than 1% of salmonellosis cases reported in the United States in the past five years (Table 1). This rarity combined with genetic, geographic, and temporal relatedness, suggests that *S.* Baildon in the wastewater was from active or asymptomatic human infections.

Our study and previous investigations suggest that wastewater-based surveillance can augment traditional patient-provider encounter-based surveillance. *S.* Derby strains isolated from wastewater in Hawaii were associated with asymptomatic cases undetected by the traditional surveillance system (27). In Texas, wastewater testing successfully identified *S.* Heidelberg, the causative agent, during and after a 2003 salmonellosis outbreak in a closed community (28). Consistent with our findings, Sahlström et al. found that *Salmonella* serotypes from sewage and salmonellosis cases in Sweden were genetically indistinguishable (29). Moreover, when Berge et al. compared *Salmonella* serotypes from two wastewater treatment plants in California with similar bacteria from patients, they concluded that unreported infections likely accounted for the differences in serovars observed (30).

The *S.* Baildon isolates we identified in wastewater in Pennsylvania were genetically indistinguishable from those epidemiologically linked to the consumption of contaminated salad. *S.* Baildon has caused two others previously reported multi-state outbreaks, which reoccurred 10 years apart starting in 1998 ((31) and (https://www.cdc.gov/salmonella/2010/restaurant-chain-a-8-4-10.html)). Traceback investigations concluded that contamination occurred on farm and in the cutting and packing facilities in the 1998-1999 and in the 2010 outbreaks, respectively. The source of the contaminated produce responsible for the *S.* Baildon outbreak that was concomitant with our wastewater detection was not identified.

The current study found that most of the genetically similar outbreak-associated *S.* Baildon cases occurred in relative proximity to the catchment areas where wastewater samples were collected. The CDC estimates that only 1 in 29 *Salmonella* infections are identified by traditional surveillance (32). Severe *Salmonella* cases including those caused by *S.* Baildon are likely reported to public health authorities ((31) and (https://www.cdc.gov/salmonella/2010/restaurant-chain-a-8-4-10.html)), whereas mild or asymptomatic infections are not (1–4). The available crude incidence rate for *Salmonella* per 100,000 was 17 cases prior to 2019 and 16.3 cases thereafter (1–2). The potential for unreported illnesses is likely because the wastewater catchment area included two facilities where residents have an elevated risk for gastroenteritis. Carceral populations experience a disproportionately high rate of foodborne infections including salmonellosis (33).

Our study was limited by the collection of samples from only two wastewater plants, both convenient to our laboratory. However, the catchment area population is comparable to those in the typical rural municipalities in Pennsylvania, which are home to about 26% of the state’s 13.0 million residents (https://www.rural.pa.gov/data/rural-quick-facts). The less than a month duration of the wastewater testing is also a limitation as isolation rates for *Salmonella* likely vary over time. Another weakness inherent in the use surveillance data is the uncertainty regarding reported cases of most infectious diseases. The use of multiple sources of data in the current study is one of its strengths. The domestic sewage we tested excluded non-human sources of *Salmonella* such as wild animals or contaminated poultry products. Although absolute certainty is impossible, the genetic association with a concurrent outbreak in the area is consist with an origin in contaminated salad. Based on FDA NARMS retail food data, *S.* Baildon has never been detected in the more than 200,000 samples tested since the inception of the program in 1997 (data not shown). Antimicrobial resistance genes are commonly found in *Salmonella* isolated from poultry meat (https://www.fda.gov/animal-veterinary/national-antimicrobial-resistance-monitoring-system/narms-now-integrated-data), but these genes were not detected in the *S.* Baildon isolated from wastewater.

Our study corroborates observations reports from investigations in Hawaii, Texas, Sweden, and California on the value of wastewater surveillance for *Salmonella* (27–30). Previous studies have cited obstacles to wastewater monitoring include complexity, perceived threat to privacy, untimeliness, insensitivity, and lack of specificity (34–35). However, the anonymity and non-intrusiveness of wastewater monitoring makes it attractive for illuminating the blind spots in the traditional surveillance systems (3–4, 36). Wastewater monitoring could be explored to augment the current resource-intensive patient encounter system in populations at elevated risk for salmonellosis outbreaks, in particular among the 1.9 million people living in over 6,000 carceral settings such as local jails, state prisons, and immigration detention facilities (33, https://www.prisonpolicy.org/reports/pie2023.html#bigpicture). Additionally, foodborne outbreaks are common among an estimated 1.3 million individuals (some frail) living in more than 1,500 predominately for-profit nursing home facilities (https://www.cdc.gov/nchs/fastats/nursing-home-care.htm, 37). Our study demonstrates that pathogen genomics could be used through partnership among public health jurisdictions and academic collaborators to monitor wastewater to enhance surveillance for *Salmonella*, particularly in vulnerable populations.

## Data availability

Sequence short reads for all wastewater isolates were uploaded to the NCBI under BioProject PRJNA357723. Accession numbers are provided in Supplementary Tables 1 and 2 for isolates from sewage and clinical sources, respectively.

## Acknowledgements

Funding was provided by the Centers for Disease Control and Prevention Epidemiology and Laboratory Capacity (ELC) for collaboration in National Antimicrobial Resistance Monitoring (CDC-RFA-CI10-101204PPHF13), U.S. Food and Drug Administration grant number 1U01FD006253-01 to PADOH for collaboration in the National Antimicrobial Resistance Monitoring System, and the US Food and Drug Administration (Grant No. 1U19FD007114-01) to E.G.D. E.M.N was funded by a USDA-NIFA Postdoctoral Fellowship 2021-67034-35120. E.G.D. was additionally supported by USDA National Institute of Food and Agriculture and Hatch Appropriations PEN4826, and J.K. was supported by PEN04853 and Multistate project 4666. No sponsors had any role in the design and conduct of the study; collection, management, analysis, or interpretation of the data; or in preparation, review, or approval of the manuscript.

